# Protocol for CHIEF (cochlear implants and inner ear inflammation) study; an observational, cross-sectional, study of children and young people undergoing cochlear implantation

**DOI:** 10.1101/2024.11.25.24317767

**Authors:** Kate Hough, Jaya Nichani, Callum Findlay, Iain A Bruce, Tracey A Newman

**Affiliations:** Clinical and Experimental Sciences, Faculty of Medicine, University of Southampton, United Kingdom; Royal Manchester Children’s Hospital, Manchester University Hospitals NHS Foundation Trust, Manchester Academic Health Science Centre, Manchester, United Kingdom; Department of Otolaryngology, University Hospital Southampton NHS Foundation Trust, Southampton, United Kingdom; Division of Infection, Immunity and Respiratory Medicine, School of Biological Sciences, Faculty of Biology, Medicine and Health, University of Manchester, Manchester, United Kingdom

**Author notes:** Corresponding author: Tracey A Newman, Faculty of Medicine, Building 85, Highfield Campus, University of Southampton, UK, SO17 1BJ, 02380599665.

**Keywords:** Inflammation, Cochlear implants, Hearing, Macrophages, Fibrosis, Transcriptome

## Abstract

**Introduction:** Cochlear implantation is a surgical treatment that restores hearing function. The implant uses a series of small electrodes to generate electrical currents in the cochlea. These currents simulate the auditory nerve to elicit hearing. Despite the success of this neuro-prosthesis, some people do not get the expected hearing benefit from their implant. One reason for this is that tissues in the cochlea vary in how they respond to the implantation of the electrode array. Many people have a healthy wound healing response that results in a mature scar tissue (fibrosis). However, some people have an altered, or heightened, inflammatory response associated with excessive fibrotic tissue growth in the cochlea. Evidence largely derived from pre-clinical studies shows that excessive tissue ensheathing the electrode array increases the electrical resistance of the current flow (impedance) and reduces the quality of electrical stimulation – both of which can lead to poorer hearing outcomes with the implant.

This study will add to our understanding of the people who have a heightened inflammatory response and more vigorous fibrosis (tissue growth) which can lead to poorer hearing outcomes. We propose that there are detectable individual inflammatory differences between people when they are implanted, which may result in variable hearing outcomes following implantation. If we could understand and identify the differences, we could detect people who may be at risk of less favourable outcomes and use targeted treatments, e.g., anti-inflammatories, to improve outcome.

**Methods and analysis:** An observational, cross-sectional, study of children and young people undergoing cochlear implantation. The study will take place at Manchester University Hospital NHS Foundation Trust (MFT) and the University of Southampton. Children and young people who meet the eligibility criteria for cochlear implantation at MFT will be invited to participate in the study. On the day of cochlear implant surgery, a sample of the middle ear mucous membrane, swabs of the nasopharynx and middle ear canal, cochlear fluid, and a blood sample will be collected. Samples will be analysed using a number of molecular techniques to determine the inflammatory status of the person at the time of implantation. Clinical hearing data will be collected for up to five years after implantation to explore the relationship between inflammation at time of implantation and long-term hearing outcomes.

**Ethics and dissemination:** This protocol has been ethically approved by IRAS (330110). Results will be submitted to international peer-reviewed journals and presented at international conferences. Results will be presented to a lay audience via our patient and public involvement and engagement group (ALL_EARS@UoS) website.

**Article Summary:** *Strengths and limitations of the study:*

- Strength: The first study that will provide the opportunity to characterise the immune state of the ear at the time of implantation and correlate it with hearing outcomes with a cochlear implant.
- Strength: The surgical protocol and the participants routine clinical care will not be altered by being a study participant.
- Strength: The first study to use spatial transcriptomics to characterise the gene expression of human middle ear macrophages in children undergoing cochlear implantation.
- Limitation: This is an observational study. The results of this study will inform sample size and recruitment criteria for future interventional studies.
- Limitation: Complete analysis and interpretation of the samples in this study requires several highly specialist and expensive techniques. This will require follow-on funding bids informed using the data from this work.

## INTRODUCTION

### Background

Cochlear implantation is a treatment developed for people with profound deafness. Cochlear implants are effective at helping young children to learn to talk and listen, older children to achieve at school, and children and young people of all ages to socialise. Children, who hear well with implants, are likely to meet key developmental milestones alongside their unaided hearing peers [1]. This contributes to a transition to adults better able to succeed in society.

Cochlear implantation involves the surgical implantation of a wire (electrode array) into the hearing organ (cochlea) in the deep part of the ear (inner ear). The electrode array carries a signal from an electronics package implanted under the skin (receiver-stimulator package), that itself receives a coded signal from a bespoke hearing aid worn on the ear (audio-processor). The electrical signals stimulate the hearing pathways in the brain, resulting in the perception of sound.

Some people do not do as well as expected following surgery or the hearing benefit from the implant tails-off over time [2–4]. This can be detrimental to the well-being of the person, result in non-use of the cochlear implant or even the need to undergo another operation to insert a new cochlear implant [5]. This can mean the potential benefits of restored hearing are lost.

How the cochlear implant electrode array interacts with the delicate tissues in the inner ear is crucial to the effectiveness of the electrical stimulation of the hearing pathway to the brain, and the preservation of any remaining natural hearing. The immune system and inflammatory response within the inner ear are likely key factors in the interaction between the electrode array and the fine structures of the ear [6–9]. We know that implantation causes an inflammatory response but most of our understanding of this comes from studies of implantation in animals [10,11] and cadaveric temporal bone studies [12,13]. Importantly these studies have not enabled us to understand how the inflammatory response might vary between people each of whom have their own immune history. We need to understand, and ideally be able to predict or anticipate, the individual inflammatory response as this could lead to and inform improved clinical management and better hearing outcomes for more people who use cochlear implants.

Our **hypothesis** is that the insertion of an electrode array causes an inflammatory response that *varies* due to individual differences between people at the time they are implanted.

We urgently need to investigate the effect that the immune state of the inner ear has on outcomes following cochlear implantation. To do this, we need to study the inflammatory state of the ear in children and young people undergoing implantation. Importantly, we need to see how this varies between children/young people, and how this is associated with hearing outcomes in people with a history of middle ear inflammation (acute otitis media and otitis media with effusion) [14,15].

The desired outcome from this study would be to understand when targeted treatment (e.g. steroids) before, during or after surgery is needed to ensure the best outcome for the person with their implant.

## METHODS AND ANALYSIS

### Study setting

The study will take place at Manchester University Hospital NHS Foundation Trust (MFT) and the University of Southampton. The Manchester paediatric cochlear implant programme was established in 1991, it serves a diverse population and implants over 60 children per year. It has a sustained record in undertaking research to improve outcomes for children and young people with their implants. The University of Southampton has significant expertise in inflammation biology, microbiology and proteomics. It is a core partner of the National Biofilm Innovation Centre and the home of one of the 19 auditory implant services in the UK (University of Southampton Auditory Implant Service).

Participant recruitment, sample collection, and all clinical management of participants will be carried out in MFT. Sample analysis and initial data interpretation will be carried out in Southampton with data sharing between centres. Only fully anonymised patient and sample data will be shared with Southampton, all identifiable information will be limited to the clinical care team in MFT.

### Study design

An observational, cross-sectional, study of children and young people undergoing cochlear implantation. Children and young people who meet the inclusion criteria and who are consented and/or give assent will be recruited to the study. The surgical protocol and the participant’s routine clinical care will not be altered by being a study participant. Figure 1 outlines the participant pathway from determining eligibility and recruitment into the study through to sample collection on the day of surgery, followed by collection of clinical and health data post-implantation.

**Figure 1.**
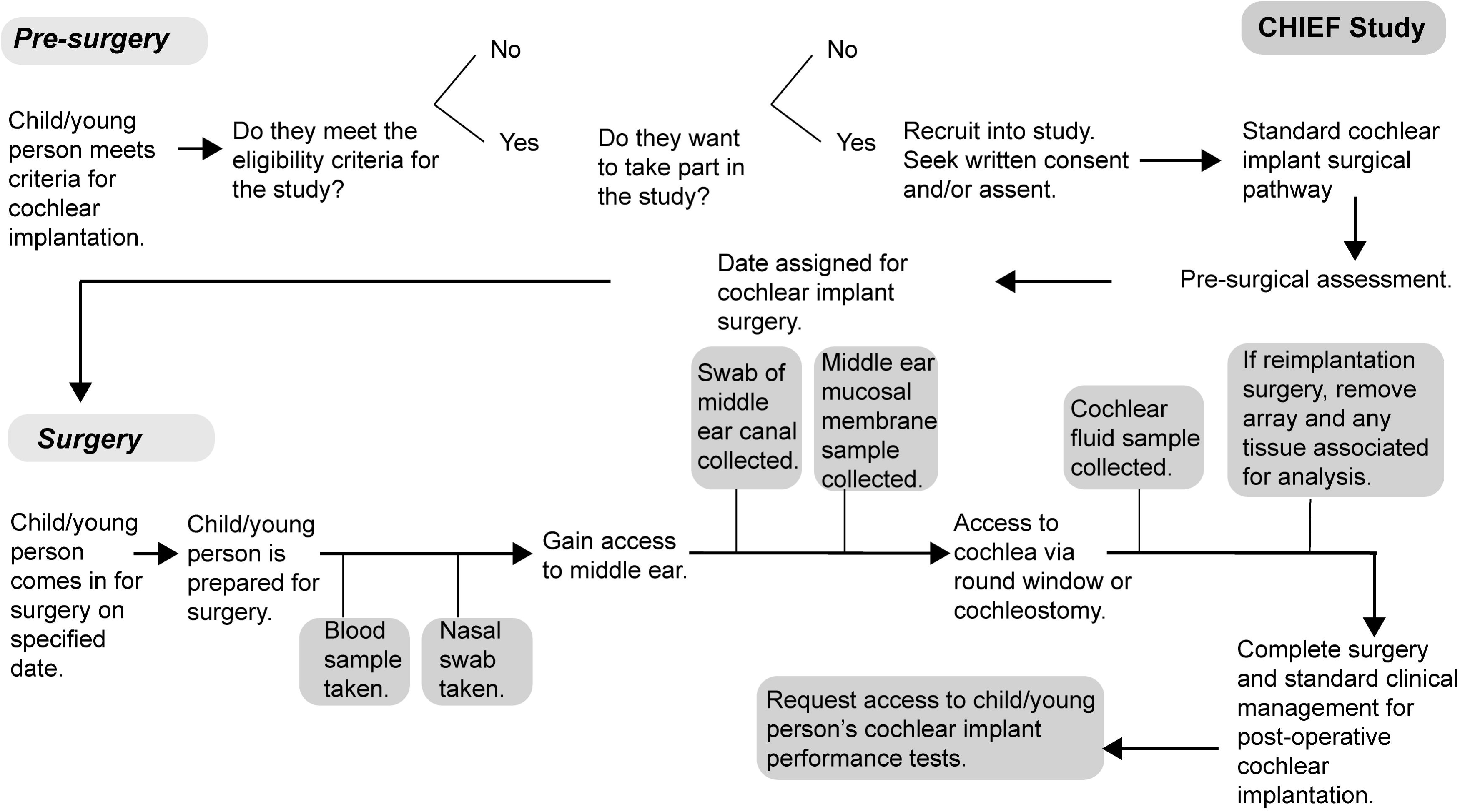
An overview of the participant pathway showing recruitment, consent, sample types and time of collection.

### Study funding

The study has been funded as part of the Manchester Hearing Health BRC award from the NIHR.

### Participants/patient recruitment

#### Eligibility criteria

Children and young people undergoing cochlear implantation under the care of the Manchester University NHS Foundation Trust (MFT) will be screened to determine if they are eligible to take part in the study. See table 1 for the study inclusion and exclusion criteria.

**Table 1.**
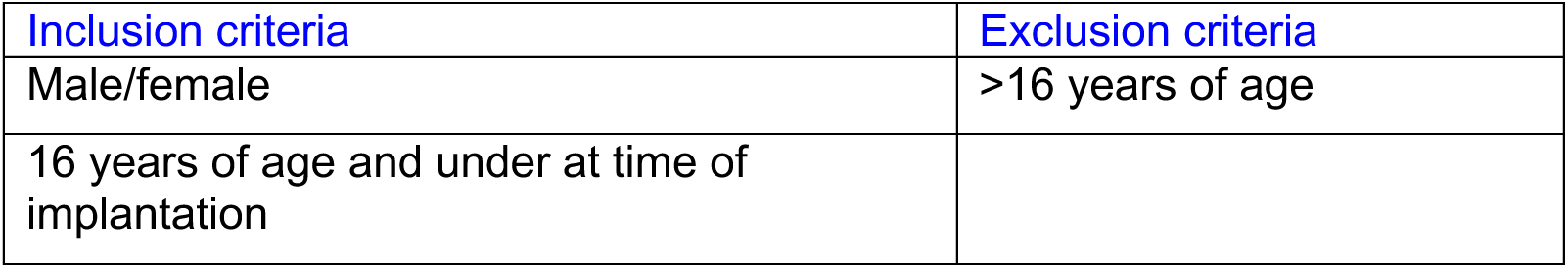
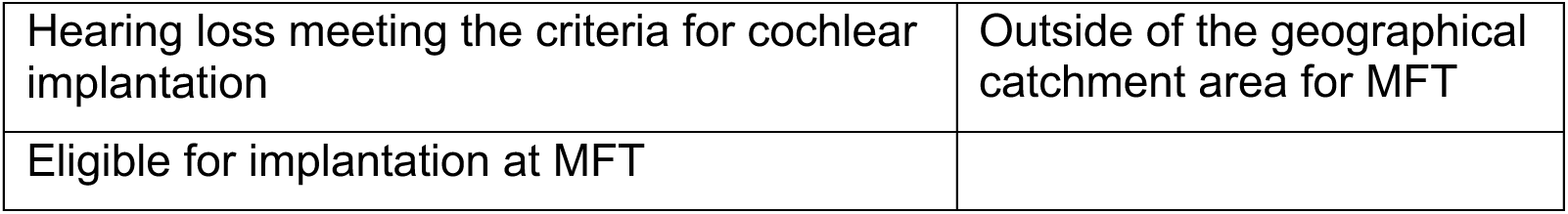
Participant inclusion and exclusion criteria.

If the patient meets the eligibility criteria, as above, the clinical care team will provide the necessary information about the study to the children and young people and/or their parents or guardians.

#### Recruitment

We aim to recruit from all children and young people who are eligible within the 24-month period of the study collection. We estimate 75 participants over the two years of recruitment to the study. This is based on recent rates of paediatric cochlear implantation at MFT.

Recruitment will be done through advertising the study to young people and the parents and guardians of children who meet the eligibility criteria. The clinical care team, with responsibility for the care of the patients, will determine eligibility. The study details will be shared during the clinic visit for those children and young people who have been identified by the clinic co-ordinator as meeting the criteria.

All eligible patients and/or their parents or guardians will receive information about the study that includes age-appropriate participant information sheets, consent and assent forms. Information about the study will be publicised on posters in the clinic, with links through quick response [QR] codes to more information about the study. The information will be hosted on our publicly available PPIE group, ALL_EARS@UoS, website [https://generic.wordpress.soton.ac.uk/all-ears/2024/11/12/chief-study-cochlear-implants-and-inner-ear-inflammation/]. The study documents have been developed with input and scrutiny of members of ALL_EARS@UoS.

#### Consent and assent

Informed consent will be obtained from a parent or guardian, and from participants aged 16 when they join the study. Informed assent forms and participant information sheets will be provided for children and young people. All participants who reach the age of 16 during the study period will be contacted and asked for their consent to continue in the study.

### Sample collection, management and storage

#### Sample collection

A sample of the middle ear mucous membranes, swabs of the nasopharynx and middle ear canal, cochlear fluid, and a blood sample will be collected at time of surgery for cochlear implantation. Samples from the ear will only be collected from the ear or ears that are being implanted.

In cases where an implant is being removed because of implant failure, together with the samples listed above we will collect any tissue attached to the implant being removed.

#### Sample management and storage

Samples will be collected and either transferred for routine analysis (blood sample - haematology), post-collection processing (tissue samples and any explanted arrays) and then storage using the biobanking facility (4°C or −80°C) at MFT.

The nasal swab, middle ear fluid, middle ear mucosa and tissue from explanted array will be stored before being transferred in batches to University of Southampton for analysis. The cochlear fluid (CF) will be stored for up to one year at MFT and transferred to Southampton at the end of year one and year two.

#### Sample processing

Middle ear mucous membrane, and tissue associated with the implant in cases being re-implanted, will be collected into pre-prepared collection tubes of 10% neutral buffered formalin [16] at surgery for tissue fixation (overnight (12-16hrs at 20-25°C), before transfer to 70% ethanol for storage (4°C) until processing. Samples will be processed to paraffin wax tissue and prepared as microarrays for histology and spatial transcriptomics.

Samples collected through swabs of the middle ear and nasopharynx will be stored before bacterial gene analysis and viral analysis. The samples will be processed to isolate the DNA using commercial kits for low (low blood contamination) and high biomass (fluid samples and where blood is present) and the 16S DNA quantity will be determined.

Cochlear fluid collected immediately prior to the insertion of the cochlear implant array will be analysed for the presence of proteins. Samples will be processed using an existing technique [17] the data will be analysed using bioinformatics.

### Sample analysis

#### Immune cell identification and characterisation from tissue analysis

##### Histological analysis

Using our published method [18], antibodies for macrophages, activated macrophages, T cells, fibroblasts, endothelial cells and a marker of cellular proliferation and appropriate counterstain will be used to identify the gross tissue morphology and distribution of cells within <4μm tissue sections. Appropriate controls will be used, and samples will be processed in batches to reduce inter-sample variation. Cell counts will be performed on each sample, with final data expressed as cells/unit area of tissue after image capture and analysis of the tissue sections using quantitative image analysis microscopy with a custom ImageJ plugin.

##### Spatial transcriptomic analysis

Gene expression and the spatial transcriptome profile at single cell resolution of immune cells (macrophages) from middle ear mucosa tissue samples will be generated in a subset of samples [19–21]. We will analyse the expression profiles to provide unbiased characterisation of the activation state and ‘memory’ of macrophages between, and within, samples using the high-plex, spatial molecular imaging platform, CosMx (manufactured by NanoString) [22]. Initial analysis and data visualisation will be done using AtoMx, a cloud-based, fully integrated spatial informatics platform.

A **primary aim** of this work is to understand whether inflammation is a factor in the inter-individual variability in the response to cochlear implantation. Macrophages are long-lived cells and have ‘a memory’ of exposure to injury or infection. This ‘memory’, known as activated or primed [6,23,24], can cause the macrophage to generate a larger inflammatory response if it is stimulated by a second injury or infection. The primary inflammatory response in macrophages is essential to drive repair, remodelling and recovery as might occur when macrophages are responding to clear an infection, or pathogens, in the inner ear. However, an over exuberant inflammatory response, in primed macrophages, will cause bystander damage to the delicate tissues of the cochlea [25,26] and may contribute to the development of scar tissue, or fibrosis, around the implant. Fibrosis around the array insulates the electrodes and alters the release of electrical current from the electrodes to their intended target for stimulation, the spiral ganglion cells of the auditory nerve. This change in electrical behaviour can be measured as increased impedance [27–29]. There is evidence that fibrosis is associated with migration of the electrode [30–32]. Altered current spread and movement in the electrode array is likely to perceived and measured as a change, or poorer, hearing outcome with the implant. The gene expression patterns, determined through bioinformatic analyses, of the macrophages will enable us to achieve our primary outcome for this part of the study which is to characterise the macrophage ‘memory’ and how this differs within and between cases. This phase of the work is a pilot study, if successful we aim to secure additional funding to characterise the response in all the study cases.

A **secondary outcome** of this phase of the work will be a dataset that captures the gene expression pattern of other cell types in the tissue. This dataset will be used in the development of follow-on studies from this work. The data will be made accessible to other researchers, with appropriate ethical approval, on request via data curation through the University of Southampton library. We will monitor and consider the most appropriate data hosting site as the project evolves. If a bespoke discipline-specific externally hosted data repository becomes available, we may store fuller data sets externally. This will be built into future funding applications.

#### Identification of bacterial species

Nasal and middle ear swabs will be analysed using culture-independent 16S ribosomal RNA (rRNA) gene amplicon sequencing to identify bacterial species within these sites. This method will enable identification of different strains of bacteria on the mucosal surface (swabs) or within middle ear fluid [33]. The samples will be processed to isolate the DNA using commercial kits for low (low blood contamination) and high biomass (fluid samples and where blood is present) and the 16S DNA quantity will be determined. Quantitative PCR will be carried out to determine the bacterial populations and the relative proportions of the populations. A known issue with using a non-culture method for the identification of bacterial strains and species is an increased likelihood of false-positive results [34], anonymised positive control samples and sample spiking with will be included within our analysis protocols to mitigate the risk of false positives.

#### Identification of viruses

Samples collected through swabs of the middle ear and nasopharynx will be stored before viral analysis.

A **key aim** is to understand how the bacterial or viral populations relate to the state of inflammation of the tissue in the middle ear in children and young people at the time of cochlear implantation. Bacteria in the middle ear have been studied in children [35] and there is some published work on bacterial analysis of adults undergoing implantation [33]. However very few children and young people undergoing implantation have been included in this work and the relationship with inflammation and hearing outcomes is not known and has not been systematically studied. This study will provide the first data of this type.

A **secondary aim** is to investigate the relationship between the microbiota of the middle ear and nasopharynx. A secondary outcome of this study is to identify whether there is a distinct middle-ear microbiota, or biomarker, that is associated with a poorer hearing outcome with a cochlear implant. This would be a significant new finding which may influence clinical management of the middle ear prior to surgery.

If the middle ear microbiota is identified as a biomarker for hearing outcomes with a cochlear implant, there would be further limitations and challenges to address as sampling the middle ear is an invasive procedure. Therefore, if during this study, we find that the nasopharynx microbiota mirrors that of the middle ear it may be that routine nasal swabs could be collected at home from children with a history of middle ear infection and sent to a laboratory for analysis. This could provide a more tolerable, easier to complete pre-surgical test and one that can be readily repeated to monitor improvements after e.g., treatment with antibiotics.

#### Identification of inflammatory markers in the blood

Blood samples will be collected on the day of surgery and analysed in the hospital haematology department to provide a full blood count on the day of surgery. The counts will be analysed, differential white blood cell count and neutrophil to leucocyte ratio, to determine the presence of systemic inflammation at the time of surgery. Previous studies, including a cross-sectional analysis, have investigated the relationship between inflammation and hearing loss [36–39]. The mechanism linking inflammation to changes in the cochlea that result in poorer sensorineural hearing loss in later life is not understood. This work will add new information about these changes in early life.

A **key aim** of this work is to determine if there is an association between levels of systemic inflammation at the time of implantation and hearing outcome following implantation. Does increased levels of systemic inflammation correlate with an increased immune/inflammatory state in the middle and inner ear and does this influence the tissue response following cochlear implantation?

#### Identification of inflammatory markers in the cochlea

Proteomics will be used to characterise the levels of inflammatory markers in the cochlea [17,40–42]. Proteomics is an unbiased technique that enables all proteins within a sample to be detected, identified and quantified. The high sensitivity of the technique enables very low levels of protein to be detected and quantified. This unbiased approach to protein detection means all proteins in the sample above the detection limits can be identified and their relative expression determined. A limitation to this can be introduced by samples that require large proteins, such as those found in blood, to be stripped from the samples prior to analysis. Low expression proteins, or protein with a high affinity for blood proteins, may result in some proteins being lost or reduced. We will use existing techniques to collect and analyse the samples [17] with specialist technical support from our proteomics and bioinformatics research units. These results will enable us to determine whether the cochlea is showing evidence of inflammation, as determined on the basis of the proteins present in the cochlear fluid [43], prior to the surgical insertion of the implant.

A **key aim** of this work is to understand whether some of the differences in response to cochlear implantation are due to individual differences in inflammation in the cochlea at the time of implantation. There are no published, or established, techniques to measure or detect these differences in the intact cochlea of people prior to implantation. However, there is evidence that there may be differences between people as in some cases people who have had meningitis, labyrinthitis and other auto-immune induced SNHL have fibrosis, or the growth of tissue, within the cochlea [44] at the time they have their cochlear implant inserted. Fibrosis occurs in many organs, such as the liver or lung [45] and importantly inflammation is a consistent feature irrespective of which organ is affected. On this basis, we predict that some people will be more inflamed and that there is need for a method to anticipate this. We will integrate the information from the cochlea at time of implantation with the data that we collect from the tissue and swab samples. Together this may enable us to start to develop a way of predicting children and young people who are at greater likelihood of inflammation in their cochlea prior to implantation or to a stronger or more prolonged inflammatory response after implantation, both of which may mean they hear less well with their implants and/or initially preserved natural low-frequency hearing is not maintained [46]. In the longer term, it may be possible to identify these children and young people for more aggressive anti-inflammatory management when they have their implant.

A secondary outcome of this phase of the work will be a data set that captures the protein expression of the fluid in the cochlea in a cohort of children and young people. This data set will be developed and used in the development of follow-on studies from this work, the data will be made accessible to other researchers on request via data curation through the University of Southampton library.

#### Clinical data collection

To explore the relationship between the immune state of the middle ear tissue and long-term hearing outcomes, we will request access to anonymised, routine clinical outcome measures for up to 5 years post-implantation including post-implantation complications, electrode impedance, deactivated electrodes, hearing and device measurements over time with the device and hearing measures. We will access data from medical records including date of birth, ethnicity, history and cause of deafness, history if ear disease (infections, surgery) and a history of other infections (meningitis, cytomegalovirus, measles).

### Research questions and objectives

Research questions:

1. What is the immune state of the middle ear in children and young people undergoing cochlear implantation?
2. How distinct or similar is the microbiota (bacteria and viruses) of the nasopharynx and middle ear, within individuals?
3. What is the relationship between the microbiota and the immune status of the middle ear tissue?
4. What is the inflammatory state of the cochlear fluid at time of surgery?
5. What is the inflammatory state of tissue associated with the cochlear implant electrode array? And how does this relate to the other sites tested in the study?
6. What is the relationship between the inflammatory state of the middle ear and outcome of cochlear implantation?

Objectives:

Use molecular techniques to;

1. identify the immune cells that are present.
2. characterise the inflammatory state of these cells, in the samples of middle ear mucous membrane (MM), or tissue attached to an implant (FT).
3. identify bacteria that are present and the size/distribution of the populations, in the middle ear (ME) fluid and nasal swab (NS).
4. identify viruses that are present and the size/distribution of the populations, in the middle ear (ME) fluid and nasal swab (NS).
5. detect the presence and levels of inflammatory markers in the cochlear fluid (CF) and blood sample (S).

Analyse clinical outcomes routinely measured across five years for the study participants.

#### Primary Outcome

The study intends to identify differences in inflammation in the ear between children and young people at the time they undergo cochlear implantation.

#### Secondary Outcome

A secondary outcome of this study is to identify the relationship between inflammatory status of the ear and the microbiota of the ear.

A secondary outcome of this study is to identify the relationship between the immune state of the ear in children and young people at the time of implantation and hearing outcomes following implantation.

### Ethics and dissemination

IRAS Number: 330110

ERGO Reference: 89599

This protocol has been ethically approved by IRAS (330110). Results will be submitted to international peer-reviewed journals and presented at international conferences. Results will be presented to a lay audience via our patient and public involvement and engagement group (ALL_EARS@UoS) website (https://generic.wordpress.soton.ac.uk/all-ears/).

### Patient and public involvement

ALL_EARS@UoS is a patient and public involvement and engagement (PPIE) group that was first established at the University of Southampton in March 2022. The group is committed to improving understanding of the mechanisms, lived experience and management of hearing loss through contributing to and influencing hearing loss research [47]. The chief investigator for this project (TN) and the postdoctoral fellow (KH), who will work on the project, have been central to ALL_EARS@UoS. The CHIEF study has been presented and discussed with the group for feedback within our regular PPIE meetings. All documents for the study were shared with members of the group for detailed feedback. One group member attended the ethical approval panel meeting alongside the chief investigator of the project. A group for young people is beginning to be co-developed to ensure young people are engaged with the project and our wider research.

## DISCUSSION

### What would the potential benefits be?

#### Improve patient outcomes

This study will allow us to combine spatial gene and protein expression data with clinical and hearing data to address the long-term aim of improving patient outcomes by identifying if there is a predictable relationship between inflammatory status and hearing outcomes following implantation.

Initially, we will determine if the inflammatory state of the middle ear, at the time of implantation, is different between children and young people using histology and spatial transcriptomics. We will combine the molecular data with clinical and hearing data to see if there is an association with hearing and surgical outcomes following implantation. This will provide essential pilot data to support further funding applications.

We hypothesise that some children and young people who have a heightened inflammatory tissue environment in the ear due to previous inflammatory insults such as recurrent, previous infections may elicit an increased inflammatory tissue response to cochlear implantation [6] resulting in increased fibrosis. This could result in poorer hearing outcomes with the implant, both in terms of initial preservation and subsequent maintenance of residual natural hearing after implantation [46,48] and worsening of the quality of electrical stimulation over time [49,50]. If we can identify patients who are at greater risk of an increased inflammatory tissue response to implantation and therefore poor hearing outcomes, we could intervene prior to, or at, surgery with existing and novel anti-inflammatory and/or -microbial therapies and ensure closer post-implantation monitoring to improve hearing outcomes following implantation.

#### Build a rich database containing clinical and medical records of children and young people who have undergone cochlear implantation alongside building a rich tissue bank

This longitudinal, observation study design will allow us to collect and bank multiple tissue and fluid samples alongside detailed participant clinical, hearing and medical data for five years. Using these data, we will produce a rich database that will allow us to ask hypothesis-driven research questions. We will use pilot data obtained from the initial histological and transcriptomic tissue analysis to apply for larger funding bids.

#### Contribute novel data sets

Our study design alongside using spatial transcriptomics, CosMx, will contribute novel data sets including the first spatial transcriptome profile of macrophages in the middle ear tissue of children undergoing cochlear implantation, as well as the transcriptome profile of other cell types including fibroblasts which are relevant for cancer and respiratory biology.

## Data Availability

Datasets from the CHIEF study will be published in the University of Southampton PURE repository.

## Acknowledgements

The authors are grateful to members of ALL_EARS@UoS PPIE group who have shared their experiences of hearing loss and cochlear implantation, contributed to discussion and provided feedback about the CHIEF study. The authors are grateful to group member, SW, who attended the ethical approval panel meeting. The authors are grateful to the Paediatric and Adolescent Cochlear Implant Team in Manchester who will be supporting this study. Thank you to the cochlear implant patients who will take part in the study.

## Data Statement

Datasets from the CHIEF study will be published in the University of Southampton PURE repository.

## Author Statement

TAN, IB, KH and JN conceived of the study. TAN and IB previously carried out a small pilot study that confirmed the feasibility of sampling of cochlear fluid during routine surgery for cochlear implantation. IB and JN have considered the timeline and feasibility and method of sample collection during the surgical process. TAN and KH have demonstrated the feasibility of the histological analysis and identified inflammation associated with tissue on an explanted device. CF has contributed to detail of the experimental design including capture and management of the data. TAN generated the first draft of the protocol and all authors have contributed to this draft.

## Funding

This work was supported by the NIHR (BRC Hearing Health award to Manchester) [NIHR203308].

## Competing Interests Statement

All authors have read and signed the BMJ policy on declaration of interests form to confirm they have no competing interests.

KH. No competing Interests

TAN. No competing interests

CF. No competing interests

IB. No competing interests

JN. No competing interests

## Notes

### Competing Interest Statement

The authors have declared no competing interest.

### Author Declarations

Integrated Research Application System gave ethical approval for this work (330110). Ethics and Research Governance Online (ERGO) of University of Southampton gave ethical approval for this work (89744.A1).

